# Face mask fit modifications that improve source control performance

**DOI:** 10.1101/2021.09.16.21263642

**Authors:** Francoise M. Blachere, Angela R. Lemons, Jayme P. Coyle, Raymond C. Derk, William G. Lindsley, Donald H. Beezhold, Karen Woodfork, Matthew G. Duling, Brenda Boutin, Theresa Boots, James R. Harris, Tim Nurkiewicz, John D. Noti

**Affiliations:** Health Effects Laboratory Division, National Institute for Occupational Safety and Health, Centers for Disease Control and Prevention, Morgantown, West Virginia, USA; Department of Physiology and Pharmacology, West Virginia University School of Medicine, Morgantown, West Virginia, USA; National Personal Protective Technology Laboratory, National Institute for Occupational Safety and Health, Centers for Disease Control and Prevention, Morgantown, West Virginia, USA; Center for Inhalation Toxicology, West Virginia University School of Medicine, Morgantown, West Virginia, USA

**Author notes:** Corresponding author: Francoise M. Blachere, MS, National Institute for Occupational Safety and Health (NIOSH), 1000 Frederick Lane, M/S 4020, Morgantown, WV 26508-5402.

**Keywords:** Face mask, fit modification, respiratory aerosols, source control

## Abstract

**BACKGROUND:** During the COVID-19 pandemic, face masks are used as source control devices to reduce the expulsion of respiratory aerosols from infected people. Modifications such as mask braces, earloop straps, knotting and tucking, and double masking have been proposed to improve mask fit. However, the data on source control are limited.

**METHODS:** The effectiveness of mask fit modifications was determined by conducting fit tests on human subjects and simulator manikins and by performing simulated coughs and exhalations using a source control measurement system.

**RESULTS:** Medical masks without modification blocked ≥56% of cough aerosols and ≥42% of exhaled aerosols. Modifying fit by crossing the earloops or placing a bracket under the mask did not increase performance, while using earloop toggles, an earloop strap, and knotting and tucking the mask increased performance. The most effective modifications for improving source control performance were double masking and using a mask brace. Placing a cloth mask over a medical mask blocked ≥85% of cough aerosols and ≥91% of exhaled aerosols. Placing a brace over a medical mask blocked ≥95% of cough aerosols and ≥99% of exhaled aerosols.

**CONCLUSION:** Fit modifications can greatly improve the performance of face masks as source control devices for respiratory aerosols.

## BACKGROUND

SARS-CoV-2 is a highly infectious respiratory virus that is primarily transmitted by respiratory aerosols and droplets emitted during activities such as talking, breathing, and coughing.^1, 2^ In the past, aerosols have been defined as airborne particles <5 µm in diameter while droplets are >5 µm, but more recent work based on aerosol physics defines aerosols as <100 µm with droplets being >100 µm.^2^ Several factors influence SARS-CoV-2 transmissibility, including particle size, a recipient’s inhalation exposure, and their susceptibility.^3^ Because symptomatic and asymptomatic individuals infected with SARS-CoV-2 can exhibit a high viral load in their respiratory fluids,^4^ the CDC recommends that unvaccinated people wear a face mask that covers the nose and mouth while indoors to reduce community transmission during the COVID-19 pandemic.^5^ Fully vaccinated individuals are also advised to wear a face mask indoors, particularly in areas of substantial or high COVID-19 transmission.^5^ Wearing a face mask to protect others from potentially infectious aerosols and droplets, called source control, has been shown to be a highly effective infection control strategy to limit the spread of COVID-19.^6, 7^ Face masks provide a physical barrier to the expulsion of both aerosols and droplets, and offer limited personal respiratory protection against aerosols that may enter through the nose and mouth.^8-10^

As a result of the COVID-19 pandemic, the consumer market has been flooded with a wide variety of face coverings and masks that vary in fit, material, and design. To provide guidance on products that are neither a medical mask (e.g., surgical masks) as per specification F2100 for providing source control nor a respirator for providing inhalation protection (e.g. N95 respirators), ASTM International developed a standard specification for barrier face coverings (F3502) to establish a baseline on mask performance using standards including submicron particulate filtration and airflow resistance (breathability).^11^ Epidemiological, clinical, and modelling studies on the community use of face masks show a significant reduction in COVID-19 transmission,^7, 12-14^ yet the data on source control performance and the level of respiratory protection are limited. Recent laboratory studies using a respiratory aerosol simulator tested several medical masks, cloth masks, and neck gaiters, and found a 40-60% reduction in the expulsion of cough aerosols.^8^ Analogous studies using a coughing (source) and breathing (recipient) simulator inside an aerosol exposure chamber demonstrated a 96% reduction in aerosol exposure to the recipient when both simulators were double masked.^9^

To limit the transmission of SARS-CoV-2, the CDC recommends wearing a multi-layer, well-fitted face mask that forms a tight seal between the face and the mask.^15^ The presence of face seal leaks enables respiratory aerosols to escape out rather than pass through the filtering materials of the mask, consequently reducing the benefits of wearing a face mask for source control. Because medical and cloth masks tend to fit more loosely than a fitted respirator^16^, recent attention has been given to modifications that improve mask fit. Aerosol exposure simulation studies by our group looking at the effect of knotting the earloops and tucking in the pleats of a medical mask or wearing a cloth mask over a medical mask (double masking) showed a significant reduction in exposure when compared to not wearing a mask or wearing a medical mask without any modification. ^9^ A publication by Clapp et al. evaluated several popular modifications and found that fitting a medical mask with either a sleeve of hosiery or multiple rubber bands, or adjusting the ear loops with either a claw-like hair clip or a 3D printed ear guard, increased particle filtration to the wearer.^17^ Utilizing the Wells-Riley equation to mathematically predict the probability of airborne disease exposure, research by Rothamer et al. demonstrated that a poorly fitting mask diminished particle filtration, but that the addition of an elastic, frame-like mask fitter improved source control performance by reducing leaks around a mask.^18^ For this study, our group evaluated various modifications that aimed to improve the fit of a medical or cloth face mask and reduce the amount of expelled aerosols during simulated coughs and exhalations.

## MATERIALS AND METHODS

### Face masks and modifications

Studies were conducted from January 2021 through April 2021. The commercially available face masks assessed in this study included two medical masks and three cloth masks of different material (polyester blend or cotton) and ply (Table 1). Tested fit modifications, also commercially available, included a mask bracket, an earloop strap, earloop toggles, and a mask brace (Table 1). Additional fit modifications included double masking, where a 3-ply cloth mask was worn over a medical face mask; crossing the earloops; and knotting & tucking, where the earloops are knotted and the mask pleats are tucked under the knotted loop. More details on the masks and fit modifications used can be found in Supplemental Figures S1 and S2.

**Table 1.** Source control devices and fit modifications evaluated during simulated cough and exhalation studies.

| *Designation* | *Product Name* | *Brand* | *Description* |
| --- | --- | --- | --- |
| **Source Control Devices** |  |  |  |
| Medical mask 1 | Disposable Protective Mask | Excellent Artisan | 3-ply medical face mask with elastic earloops & adjustable metal nose strip |
| Medical mask 2 | Disposable Surgical Mask | Winner Medical Co., Ltd. | 3-ply medical face mask with elastic earloops & adjustable metal nose strip |
| 2-ply cloth mask | Reusable 2-ply Face Mask | Lefty Production, Co. | 2-ply polyester blend face mask with earloops |
| 3-ply cloth mask | Defender 3-ply Cotton Mask | HanesBrands | 3-ply 100% cotton face mask with earloops & adjustable metal nose strip |
| 4-ply cloth mask | Reusable 4-ply Face Mask | Badger-Smith | 4-ply cotton-polyester blend face mask with earloops & adjustable metal nose strip |
| **Fit Modifications** |  |  |  |
| Earloops crossed |  |  | Crossing the earloops to create a loop that fits over the ear |
| Mask bracket | Cool Protection Stand 3D Mask Bracket | Anbirong | Reusable plastic mask bracket worn under a face mask |
| Earloop strap | Adjustable Mask Ear Strap Hook Extender | Maoxing Weiye | Reusable plastic adjustable strap worn behind the head to adjust earloops |
| Earloop toggle | Silicone Elastic Mask Adjustment Buckle | Beeager | Silicone toggles used to adjust the earloops |
| Knotted & tucked |  |  | Knotting the earloops near the mask panel with excess material tucked under the knot |
| Double masking |  |  | A 3-ply 100% cotton cloth mask worn over a medical face mask |
| Mask brace | Mask brace | Fix the Mask | Reusable elastic brace worn over a mask |

### Filtration efficiency and inhalation airflow resistance

ASTM standardized tests for filtration efficiency and inhalation airflow resistance measurements from unmodified medical and cloth face masks were performed using automated filter testers (Models 8130 and 8130A, TSI) as previously described.^10^ Briefly, medical and cloth face masks were secured to a test plate using beeswax. Pleats on the unmodified medical face masks were expanded prior to measurement. Filtration efficiency and airflow resistance were measured on the double-mask modification by securing the edges of a 3-ply cloth mask over the edges of a medical face mask with beeswax. Filtration efficiency and airflow resistance were not measured on face masks with crossed earloops, mask brackets, earloop straps, earloop toggles, knotted and tucked earloops, or mask braces, as these fit modifications do not alter the performance of the materials used in the construction of a mask.

### Fit testing

Fit factor assessment, which measures the degree to which aerosols can enter through face seal leaks on a mask, was performed using the PortaCount Pro+ respirator fit tester as previously described.^10^ Reported fit values are reflective of the ratio of the aerosol concentration outside the face mask to the aerosol concentration inside the face mask.^10^ For fit tests on human subjects, a sample group of 4 subjects participated in the study. Face masks with and without modification were fit tested on human subjects (n=3-4 tests/modification) with a PortaCount® Pro+ (model 8038, TSI Corporation; Shoreview, MN) in N99 mode using the Occupational Safety and Health Administration (OSHA) modified ambient aerosol condensation nuclei counter (CNC) protocol for filtering facepiece respirators.^19^ Because only fit factors were measured and no identifiable information was collected, the West Virginia University Office of Human Research Protections determined that Institutional Review Board approval was not required for this study. This activity was reviewed by CDC and was conducted consistent with applicable federal law and CDC policy (see e.g., 45 C.F.R. part 46; 21 C.F.R. part 56; 42 U.S.C. §241(d), 5 U.S.C. §552a, 44 U.S.C. §3501 et seq.). Simulator manikins were fit tested with each mask fit modification (n=4-5 tests/modification) using a PortaCount® Pro+ (Model 8038, TSI Corporation; Shoreview, MN) in N99 mode as per manufacturer’s instructions. A constant breathing rate of 36 L/min was used for all simulator fit tests and a daily quality assurance test was conducted using a 3M 1860 N95 respirator.

### Source control measurement

A source control measurement system (Supplemental Figure S3) was used as previously described to measure the collection efficiencies (% particles blocked) for coughed or exhaled aerosols by fit modified or unmodified face masks.^8, 10^ Briefly, a test aerosol solution consisting of 14% potassium chloride (KCl) and 0.4% sodium fluorescein (particle size range from 0-20 μm in diameter) was propelled through the mouth of an elastomeric headform outfitted with a mask (with modification or without modification) during simulated coughs (4.2 L volume) and breathing (15 L/min) into a 136 L collection chamber. Each face mask (unmodified or modified) was used for two consecutive tests with a total of four experimental replicates performed under the set experimental conditions for simulated cough and exhalations. The performance of a 3M 1860 N95 respirator as a source control device was also included in this study for metric comparisons. The test aerosol collected from control experiments without a mask had a total mass of 525 μg (cough) and 495 μg (exhalation). An Anderson Impactor operating at 28.3 L/min was used to collect and separate the test aerosol into seven particle size fractions by their aerodynamic diameter: <0.6 μm; 0.6-1.1 μm; 1.1-2.1 μm; 2.1-3.3 μm; 3.3-4.7 μm, 4.7-7.0 μm; and >7 μm. Because of possible losses from settling, particle data for the >7 μm size fraction (<0.7% of the total test aerosol mass) was not included in the collection analysis.

### Exposure reduction studies

Respiratory exposure studies were performed using a simulator that expels a test aerosol (the source) and a breathing simulator (the recipient) inside an experimental chamber as previously described.^9, 20^ Briefly, a medical face mask or cloth mask (with or without fit modification) was placed on the source simulator situated 6 feet from the recipient simulator. An optical particle counter (Grimm 1.108; Aerosol Technik Ainring GmbH & Co. KG; Ainring, Germany) was used to measure the aerosol concentration at the mouth of an unmasked recipient simulator. Exposure was assessed by comparing the mean mass aerosol concentration measured at the mouth of the unmasked recipient when no mask was worn by the source simulator compared when a fit modified medical or cloth mask was worn.

### Statistical analysis

Mask source control performance was assessed by calculating the collection efficiency as (1 – M_mask_/M_control_), where M_mask_ = total mass of the aerosol particles that passed through or around the fit modified source control device and was collected by the impactor and M_control_ = total mass of the aerosol particles expelled by the source control measurement system without a face mask and collected by the impactor. To test for significance for cloth mask types that only had one type of modification versus the control, the Wilcoxon rank sum test was used. For mask types that contained more than 1 level of fit modification, overall significance was first assessed using the Kruskal-Wallis test. Pairwise comparisons were then made using the Wilcoxon rank sum test and p-values were adjusted using the Benjamini, Hochberg, and Youkilis method to control the false discovery rate to compare each fit modification to the unmodified mask control. The percent change in collection efficiency was considered significant if p ≤ 0.05. Fit factor, filtration efficiency, and inhalation airflow resistance data were analyzed using a pairwise Wilcoxon rank sum test followed by a Benjamini-Hochberg correction for multiple comparisons. Each fit modification method was compared to the unmodified mask control. Differences were considered significant at a p ≤ 0.05. All statistical analyses were performed using R Statistical Environment v. 4.0.2 (The R Foundation, Vienna, Austria).

## RESULTS

### Filtration efficiencies and inhalation airflow resistance

Filtration efficiency and inhalation airflow resistance measurements show differences between the material and ply level of medical and cloth face masks (Supplemental Table ST1). Medical mask 1 had a filtration efficiency of 82.0% and an airflow resistance of 45.4 Pa whereas medical mask 2 had a filtration efficiency of 96.4% and an airflow resistance of 63.7 Pa. The combination of a 3-ply cloth mask over medical mask 1 demonstrated a similar filtration efficiency of 83.3%, but airflow resistance increased to 98.7 Pa. Likewise, medical mask 2 doubled with a 3-ply cloth mask had a filtration efficiency of 95.5% but airflow resistance increased to 97.1 Pa. The filtration performance for cloth face masks was 2 to 4-fold lower than that of the medical masks and airflow resistance was generally higher. The 2-ply cloth mask exhibited a filtration efficiency of 20.2% and an airflow resistance of 96.4 Pa while the 3-ply mask had a filtration efficiency of 21.0% and an airflow resistance 45.1 Pa. The 4-ply cloth mask had an elevated filtration efficiency of 36.0% and an airflow resistance of 92.2 Pa.

### Human and manikin fit tests

To assess mask fit with and without modification, quantitative fit testing was performed on human subjects and on the simulator manikin (Table 2). The average human fit factor for medical mask 1 was 1.6, while medical mask 2 had an average fit factor of 1.8; these results are consistent with previous studies that reported human fit factors of 1.0 to 2.4 for medical and cloth face masks.^10^ Crossing the earloops or using a mask bracket decreased the fit factor of both medical masks. The remaining fit modifications increased the fit factor, with the mask brace demonstrating the greatest increase to 7.2 for medical mask 1 and 13.3 for medical mask 2. Increases in fit factor were also observed when the mask brace was secured over a cloth mask, with the 4-ply cloth mask demonstrating a 3-fold increase in fit factor.

**Table 2.** Human and manikin mask fit factors evaluated during mask fit tests using a PortaCount^®^ Pro+ (TSI).

| **Face Covering** | **Modification** | **Human Fit Factor** |  | **Manikin Fit Factor** |  |
| --- | --- | --- | --- | --- | --- |
|  |  | **mean** | **SD** | **mean** | **SD** |
| Medical mask 1 | No modification | 1.6 | 0.5 | 2.0 | 0.2 |
|  | Crossed | 1.1 | 0.2 | 2.2 | 0.4 |
|  | Bracket | 1.0 | 0.0 | 2.3 | 0.1 |
|  | Strap | 5.4 | 3.2 | 2.9 | 0.4 |
|  | Toggle | 4.0 | 2.0 | 4.1 | 0.6 |
|  | Knotted & tucked | 6.0 | 1.5 | 4.0 | 0.9 |
|  | Double mask | 4.2 | 2.6 | 6.7 | 2.0 |
|  | Brace | 7.2 | 1.0 | 6.1 | 2.0 |
| Medical mask 2 | No modification | 1.8 | 0.4 | 2.9 | 0.7 |
|  | Crossed | 1.1 | 0.2 | 2.3 | 0.6 |
|  | Bracket | 1.6 | 0.5 | 2.2 | 0.1 |
|  | Strap | 3.3 | 1.2 | 3.1 | 0.6 |
|  | Toggle | 6.0 | 5.8 | 5.3 | 1.6 |
|  | Knotted & tucked | 6.3 | 3.6 | 4.8 | 0.7 |
|  | Double mask | 2.1 | 1.1 | 7.0 | 2.9 |
|  | Brace | 13.3 | 3.7 | 7.2 | 1.3 |
| 2-ply cloth mask | No modification | 1.4 | 0.3 | 1.5 | 0.0 |
|  | Brace | 2.0† | 0.0 | 2.0 | 0.3 |
| 3-ply cloth mask | No modification | 1.3 | 0.5 | 1.5 | 0.1 |
|  | Brace | 2.0 | 0.0 | 2.1 | 0.3 |
| 4-ply cloth mask | No modification | 1.5 | 0.6 | 2.6 | 0.5 |
|  | Brace | 4.6† | 1.5 | 3.6 | 0.7 |

†Fit factor was determined to be statistically significant ( $p < 0.05$ ) when comparing a fit modified face mask to the corresponding no modification control.

When fitted with a medical mask, the average manikin fit factor was 2.0 for medical mask 1 and 2.9 for medical mask 2 (Table 2). As was observed in human fit testing, crossing the earloops decreased the fit factor for medical mask 2 on the manikin. Some modifications, such as double masking and donning a mask brace, demonstrated a 2.4 to 3.3-fold increase in mask fit. An increase in mask fit was also observed when the mask brace was secured over a cloth mask, with the 3-ply and 4-ply cloth masks demonstrating a 1.4-fold increase in fit factor.

### Source control simulation studies

Using the source control measurement system, the efficacies of unmodified and fit modified face masks at collecting aerosol particles expelled during simulated coughs and exhalations are presented in Figure 1. The mean particle collection efficiency of medical mask 1 without modification was 56.0% for coughing and 42.0% for exhalations, while medical mask 2 had a collection efficiency of 63.0% for coughing and 55.0% for exhalation. Crossing the earloops on a medical face mask or using a mask bracket did not significantly improve the source control performance. Increasing tension to the earloops with an adjustable strap significantly improved the collection efficiencies of both medical masks during cough experiments to 72.0% for medical mask 1 and 75.0% for medical mask 2. The strap modification also significantly improved the collection efficiency of medical mask 1 during simulated exhalation but did not change the performance of medical mask 2. Modifications to the earloops by adding toggles or by knotting and tucking achieved similar increases in source control performance to greater than 74% for both medical masks tested. Fit modifications that produced the most significant improvement to the source control performance of a medical face mask were double masking with a cloth mask over the medical mask and the use of a mask brace. Particle collection efficiencies for double masking with medical mask 1 were 85.0% during experimental coughs and 92% during exhalation, while doubling masking with medical mask 2 demonstrated collection efficiencies upward of 92% (cough) and 91% (exhalation). When using a mask brace over medical mask 1 or medical mask 2, average collection efficiencies of 95% for coughs and 99% for exhalations were obtained.

**Figure 1.**
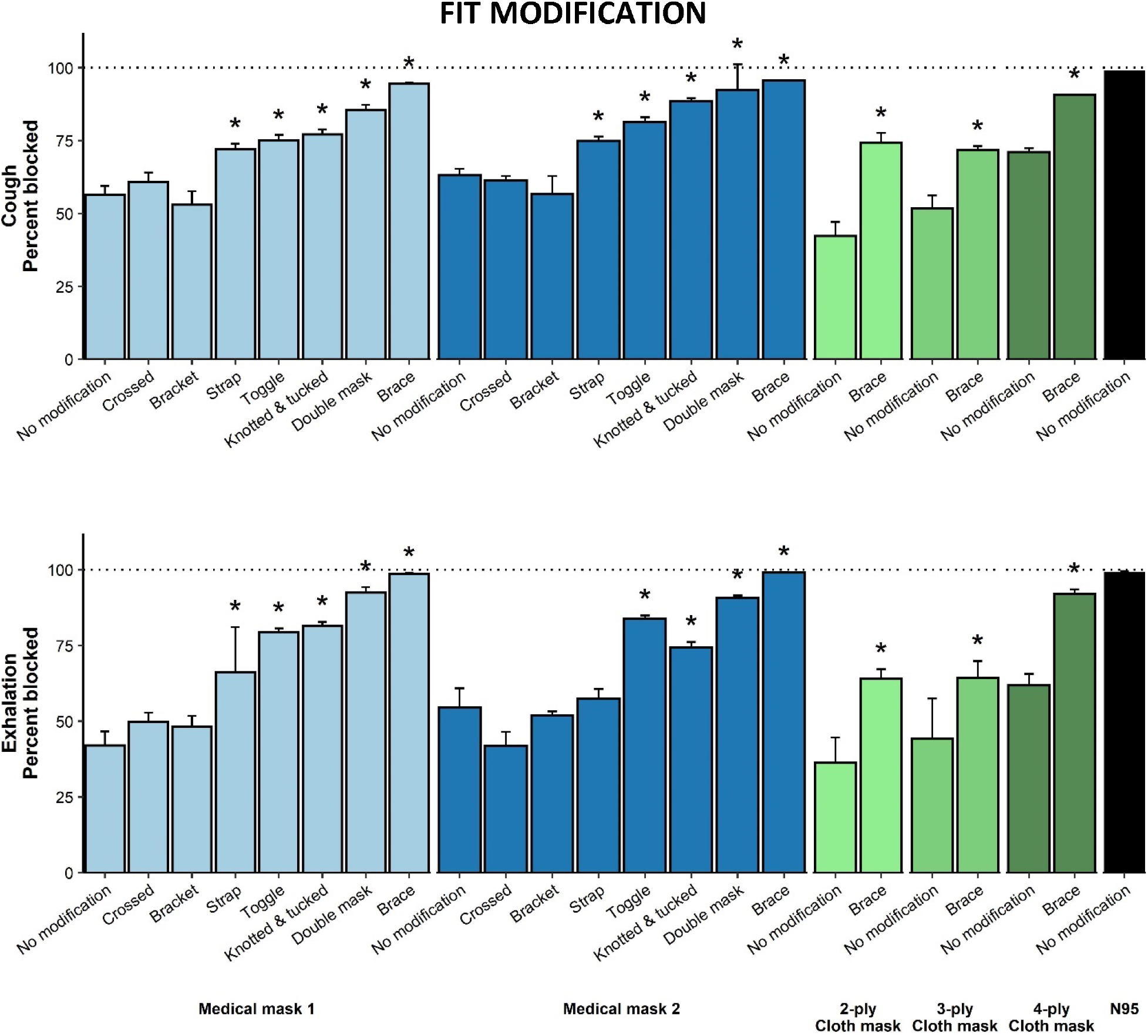
Source control performance of face masks (denoted by colors) with and without fit modifications. For comparison, source control data for an N95 respirator was included. Total particles blocked (%) by medical and cloth face masks with and without fit modifications following cough (top) and exhalation (bottom) simulations. Percentage blocked is based on mass of particles collected following unmasked source coughing and exhalation experiments. Asterisks (*) indicate the modification was determined to be statistically significant (p < 0.05) compared to the corresponding no modification control.

Respiratory viruses are transmitted by droplets and aerosols in a broad range of particle sizes.^13^ Reduced source control performance with an unmodified medical mask was largely due to a lower collection efficiency for particles ≤3.3 µm in size. Medical face masks blocked 53-60% of expelled particles ≤3.3 μm during simulated coughs but blocked upwards of 80% of the particles sized >3.3 μm (Figure 2). Similar trends were observed following simulated exhalation experiments with 40-54% of particles ≤3.3 μm and 62-71% of particles >3.3 μm collected. Fit modifying a medical mask by crossing the earloops, adding a bracket, or using an earloop strap did not increase the collection efficiency of particles in either size range. However, the other fit modifications improved the collection efficiency of particles, particularly those in the ≤3.3 μm range. The most marked improvement for particles ≤3.3 μm was observed following addition of the mask brace, where a collection efficiency of nearly 100% was obtained by securing either type of medical mask with the elastic brace.

**Figure 2.**
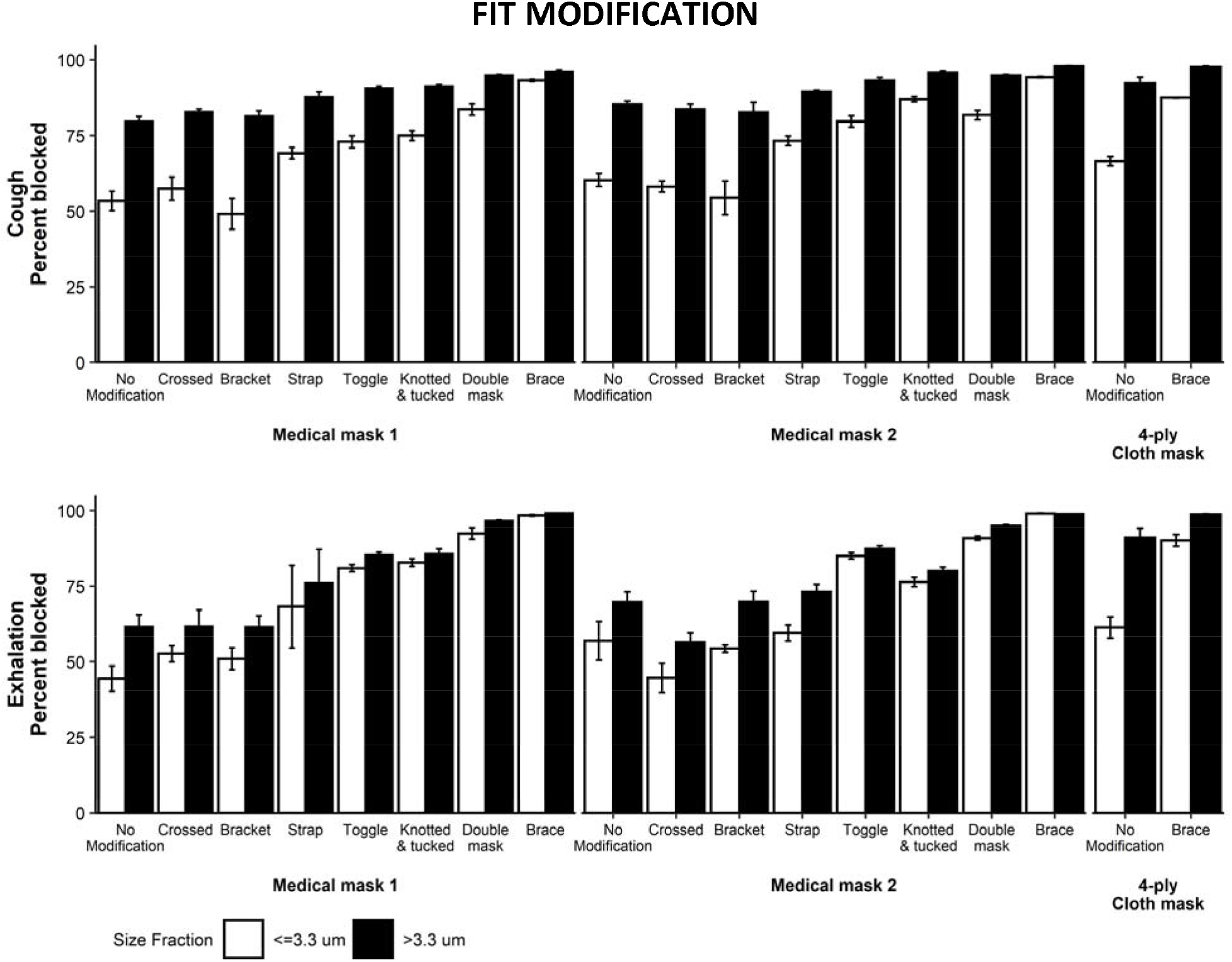
Percentage of particles in size fractions less and greater than 3.3 μm blocked by face masks with and without fit modifications. Particles blocked (%) in the size fraction ≤ 3.3 μm (white bars) and the size fraction > 3.3 μm (black bars) following cough (top) and exhalation (bottom) simulations.

Source control performance of fit modified cloth face masks with a mask brace demonstrated significant improvements in particle collection during simulated coughs and exhalations (Figure 1). The collection efficiency of an unmodified cloth mask tested during cough simulations ranged from 42% for the 2-ply cloth mask to 51% for the 3-ply cloth mask and 71% for a 4-ply cloth mask. Similarly, increasing the number of cloth layers increased the collection efficiency during simulated exhalation, with values of 36% for the 2-ply cloth mask, 44% for the 3-ply cloth mask, and 62% for the 4-ply cloth mask. Securing a 4-ply cloth mask with a mask brace further increased the collection efficiency from 71% to 91% during simulated coughs, and from 62% to 92% during simulated exhalation. An increase in collection efficiency was also observed when securing the mask brace over the 3-ply cloth mask.

When looking at the collection efficiency of cloth masks for particles ≤3.3 µm in size, neither a 2-ply nor 3-ply cloth mask performed to the level of a 4-ply mask (results not shown). The percentage of particles blocked by a 4-ply cloth mask are presented in Figure 2. Results were similar to what was observed following fit modification of a medical mask with a mask brace. The addition of a mask brace over the 4-ply cloth mask demonstrated a marked improvement when collecting particles ≤3.3 μm in size, with up to 88% and 90% blocked during simulated coughs and exhalation, respectively. For particles sized >3.3 μm, the mask brace increased the collection efficiency of a 4-ply cloth mask from 92% to 98% during simulated coughs, and from 91% to 99% during simulated exhalation.

### Exposure reduction studies

Simulated particle exposure studies examining the performance of fit modified medical and cloth face masks are presented in Figure 3. In these studies, particles were exhaled by a masked source simulator and the mass of the particles reaching the mouth of an unmasked recipient breathing simulator was measured. The reduction in exposure to airborne particles seen in these experiments were similar to the particle collection efficiencies measured with the source control system. Likewise, significant increases in particle blocking were obtained when the source simulator wore a medical mask fit modified with earloop toggles, knotted and tucked earloops, or a mask brace. Results show that greater than 98% of the particle mass was blocked by a medical face mask fit modified with a brace and are comparable to the source control performance of an N95 respirator.^8^ While the mask brace appeared to improve the percentage of particles blocked by a 2-ply and 3-ply cloth mask, the increase was not statistically significant.

**Figure 3.**
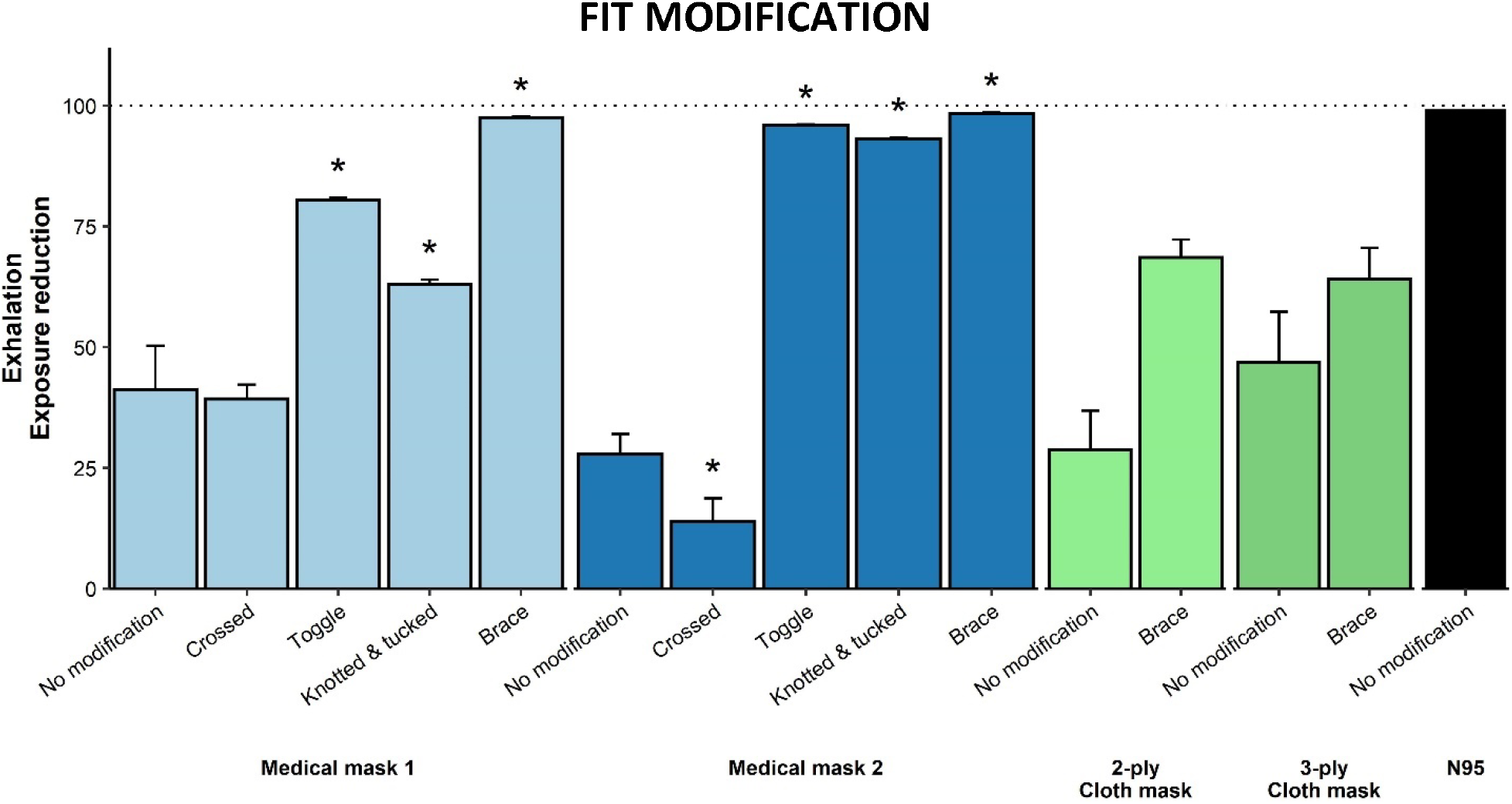
Exposure reduction (%) resulting from source masking (denoted by colors) in environmental chamber exhalation studies using both source and recipient respiratory simulators. Percent reduction is based on recipient exposures following unmasked source exhalation simulations. For comparison, expsoure reduction data for an N95 respirator was included. Asterisks (*) indicate the modification was determined to be statistically significant (p < 0.05) compared to the corresponding no modification control.

## DISCUSSION

SARS-CoV-2 is transmissible through expelled respiratory droplets and aerosols. Although the relative contribution of droplets and aerosols to COVID-19 cases remains unclear, the use of face masks is an important source control measure that reduces the expulsion of these respiratory droplets and aerosols and helps slow the spread of SARS-CoV-2.^21^ Over the course of the COVID-19 pandemic, several modifications have been suggested by media outlets to improve the comfort or fit of face masks, but little information is available as to how these fit modifications affect the source control performance of face masks. Although fit testing is an OSHA requirement used for assessing seal leakages on tight fitting respiratory protective devices such as N95 respirators and filtering facepiece respirators,^19^ fit tests of medical masks show that crossing the earloops or using a mask bracket generally diminishes the fit factor on both the simulator manikin as well as on human subjects. On the other hand, use of earloop toggles or a mask brace created a better seal and improved the fit of medical masks. The mask brace also significantly improved the fit of cloth face masks on human subjects. In general, greater fit factors were observed with the pliable headform used in our respiratory simulation studies compared to human subjects. These noted differences are consistent with previous studies by our group and likely relate to the fit test protocol and facial variations that alter how well a mask seals to the face.^10^ When measuring mask fit factor, human test subjects performed the series of test exercises outlined by the CNC fit test protocol^19^ whereas the manikin simulators used in this study are static and also breathed at a constant rate.

Using the source control measurement system, our quantitative lab-based studies demonstrate that not all fit modifications improve mask source control performance. While advertised to create more breathing space, the use of a mask bracket with a medical mask was found to reduce the total mass of particles collected compared with an unmodified medical mask. Visible face seal gaps along the cheeks of the manikin were evident when the mask bracket was inserted under a medical face mask (Supplemental Figure S1), enabling expelled aerosols to flow into the collection chamber. In comparison, a strap, toggles, or knotted and tucked fit modifications created a better mask seal to the simulator headform and enhanced the source control performance of a medical face mask. Likewise, double masking with a 3-ply cloth mask or use of a mask brace over a medical face mask created a tighter seal and significantly improved particle collection efficiencies.

Although the source control measurement system is unable to separate out the effects of multiple variables within each mask type, simulation studies examining the performance of unmodified and fit modified face masks demonstrate that fabric composition and ply level greatly affect source control performance. A greater percentage of expelled particles were blocked by an unmodified 4-ply cloth mask in comparison to an unmodified 3-ply cloth mask. These results support previous findings by our group examining performance metrics for cloth face masks as source control devices.^10^ In this current study, we found that an unmodified 4-ply cloth mask had a higher particle collection efficiency compared with an unmodified medical face mask, but a brace-modified 4-ply cloth mask did not perform to the level of a brace-modified medical face mask. The presence of seal leaks as a result of poor mask fit is a likely explanation for this discrepancy. Results from our simulation studies examining the double masking modification, where a 3-ply cloth mask was layered over a medical face mask, show that collection efficiencies increased when a tighter seal was attained on a medical mask. Consequently, layering two loose fitting medical masks would neither reduce seal leaks nor improve source control performance.

Filtration efficiency studies examining the intrinsic properties of the materials used in cloth face masks have shown that fabrics with tight weaves and low porosity, and the use of multiple layers and fabric combinations (cotton polyester blends), can effectively filter aerosol particles in the 10 nm to 10 μm size range.^22^ While mask filtration efficiency as well as airflow resistance measurements are neither reflective of mask fit nor source control performance, our study results provide supporting evidence that the materials used in the cloth mask do not filter particles to the level of the materials used in a medical face mask. When looking at inhalation airflow resistance, the assessed 2-ply and 4-ply cloth masks demonstrated elevated values in comparison to the 3-ply cloth mask or medical masks tested in this study. Likewise, layering a 3-ply cloth mask over a medical mask (double masking) noticeably increased airflow resistance. Airflow resistance, which is an ASTM standardized test that measures how breathable a mask is, has important implications with regards to user compliance. It should be noted that differences in filtration efficiency and airflow resistance were evident between the two medical face masks tested. Despite both masks consisting of 3 plies of material, medical mask 2 had a greater filtration efficiency and airflow resistance than medical mask 1 and performed better overall as a source control device when tested unmodified or fit modified. The materials used in the construction of medical mask 2 were more rigid and lent towards better facial contouring and, thus, source control performance, most notably when either toggles or the knotted and tucked ear loop modification were used to enhance the seal of the mask. Collectively, these empirical studies emphasize the importance of wearing a comfortable face mask that effectively filters respiratory particles and seals tightly to the face for optimal source control.

Humans continuously expel respiratory droplets and aerosols in a broad range of aerodynamic particles sizes. A recently published study has shown that larger respiratory droplets can travel up to 8 meters from an infected individual before settling onto surfaces, whereas smaller respiratory aerosols can remain airborne almost indefinitely.^23^ Limiting the expulsion of respiratory aerosols of any size from an infected source is critical for transmission control. Our respiratory simulator expels a test aerosol predominantly consisting of particles ≤7 μm in size. Using two different methods of aerosol measurement, our simulation studies demonstrate that not all medical and cloth masks perform equally as source control devices. During cough simulations, as particle size increased above 3.3 μm, comparable collection efficiencies were observed between an unmodified and a fit modified face mask (cloth or medical), suggesting that larger particles were more likely to be filtered out by the mask compared with those under 3.3 μm. However, when looking at the smaller size fractions of the test aerosol produced during simulated coughs and exhalations, a shift in particle size distribution and collection efficiency was evident when a face mask was fit modified. When the fit of a medical face mask was modified with a brace or layered with a cloth mask (double masking), the collection efficiency improved for particles ≤3.3 μm in size. A similar trend was observed when a cloth mask was secured with a brace. Because expelled respiratory particles are influenced by air flow dynamics, using a fit modification that reduces facial gaps along the nose and contours of the face is key for effective source control. However, additional aerosol simulation studies looking further at aerodynamic particle size distributions are warranted.

Several limitations exist with our study looking at the effect of masks and fit modifications on source control performance. Many different types of face masks are available for purchase. We tested three different cloth masks that varied in fabric composition and ply level and tested two different 3-ply medical masks composed of synthetic material of unknown formulation. Likewise, we tested seven different fit modifications and compared particle collection efficiencies with the equivalent unmodified face mask. As such, mask production-related inconsistencies may create minor discrepancies in the comparative analysis. Other limitations to our study entail the respiratory conditions used on the source control system. For our cough simulation studies, we used a single cough flow profile that is based upon earlier studies that assessed cough volumes and flow rates from influenza patients.^24^ For the simulated exhalation studies, the International Organization for Standardization (ISO) standard for the ventilation rate for a female performing light work was selected.^8^ Coughing and breathing flow rates vary from person to person under different physiological conditions, and different flow rates could give different results. The composition of the test aerosol used in our simulation studies is not comparable to human respiratory aerosols nor is the test aerosol enveloped by a turbulent gas cloud that is typically generated during human expiratory events.^25^ Lastly, results from our experimental studies assume that the fluorescein dye used in aerosol particle quantification is homogenously distributed to KCl particles.

## CONCLUSIONS

Our respiratory simulation studies examined the source control performance of medical and cloth face masks in conjunction with several fit modifications and identified practical combinations that improved mask seal and efficaciously blocked expelled aerosols from the source. Layering a 3-ply cloth mask over a medical mask (double masking) or securing a medical mask with an elastic brace improved fit and provided optimal source control performance. Further evaluation of mask source control performance through regression analysis would prove informative. The results of these studies have broad applicability towards personal measures that can be taken to reduce the transmission of respirable infectious pathogens and are not limited to the SARS-CoV-2 virus.

## Data Availability

Data is available from research personnel upon request.

## DISCLAIMERS

The authors declare no competing interests. The findings and conclusions in this report are those of the authors and do not necessarily represent the official position of the National Institute for Occupational Safety and Health, Centers for Disease Control and Prevention. Mention of any company or product does not constitute endorsement by the National Institute for Occupational Safety and Health, Centers for Disease Control and Prevention.

## ACKNOWLEDGEMENTS

We would like to thank our families for their support. This work was supported by the following sources: Centers for Disease Control and Prevention, National Institutes of Health R01 ES015022 (TRN) and WV-CTSI U54 GM104942-05.

