## Supplemental material for "Face mask fit modifications that improve source control performance"

Corresponding author:

Francoise M. Blachere, MS

| ***Face Covering*** | ***Modification*** | ***Filtration Efficiency (%)*** | | ***Inhalation Airflow Resistance (Pa)*** | |
| --- | --- | --- | --- | --- | --- |
|  |  | ***mean*** | ***SD*** | ***mean*** | ***SD*** |
| Medical mask 1 | No modification | 82.0 | 0.8 | 45.4 | 3.2 |
|  | Double mask | 83.3 | 2.4 | 98.7 | 5.4 |
| Medical mask 2 | No modification | 96.4 | 0.1 | 63.7 | 2.9 |
|  | Double mask | 95.5 | 0.9 | 97.1 | 3.5 |
| 2-ply cloth mask | No modification | 20.2 | 0.8 | 96.4 | 5.9 |
| 3-ply cloth mask | No modification | 21.0 | 2.5 | 45.1 | 1.0 |
| 4-ply cloth mask | No modification | 36.0 | 9.9 | 92.2 | 1.0 |

Supplemental Table ST1. Mask filtration efficiency and inhalation airflow resistance measurements.

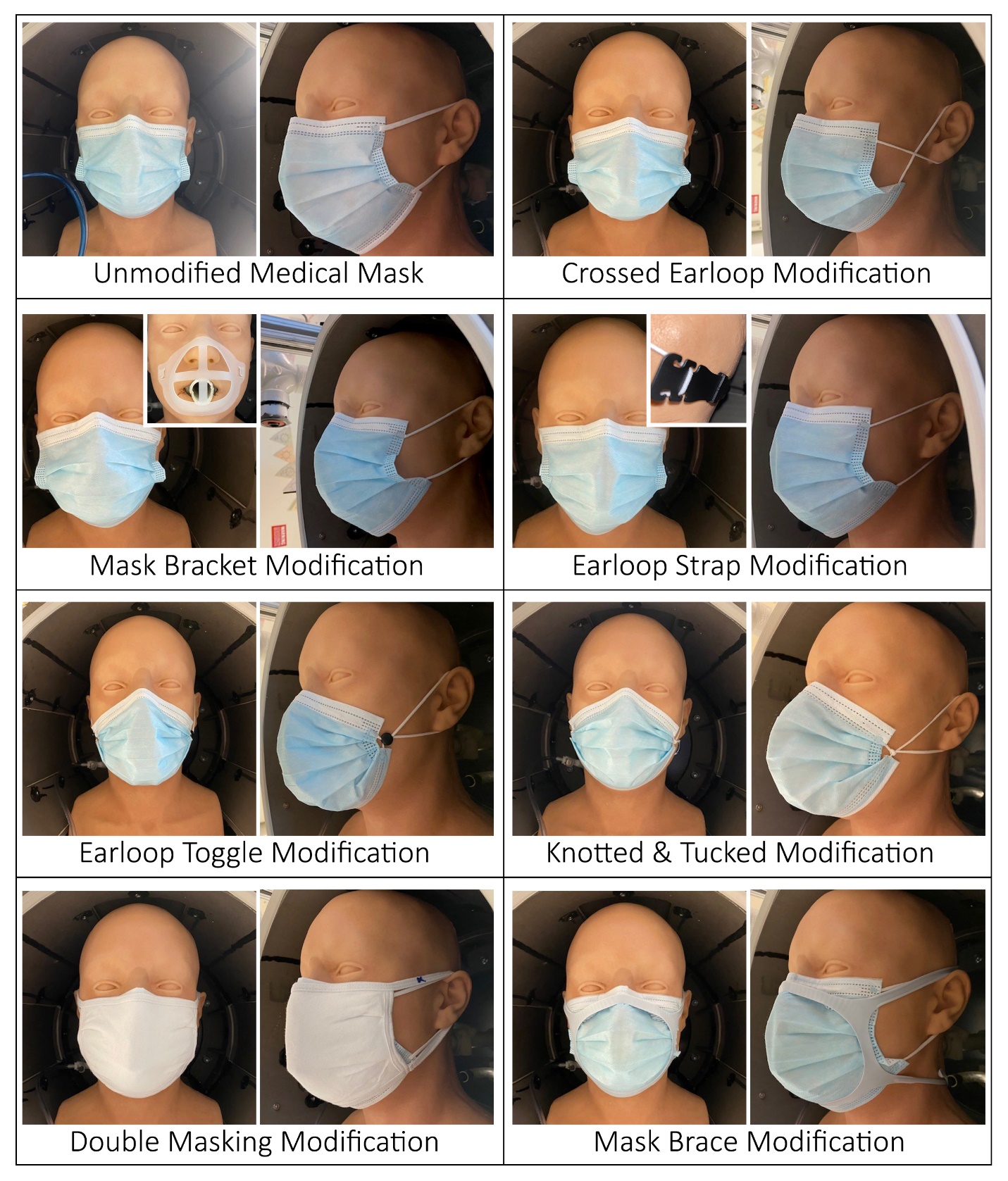

Supplemental Figure S1. Medical mask fit modifications evaluated during simulated cough and exhalation studies.

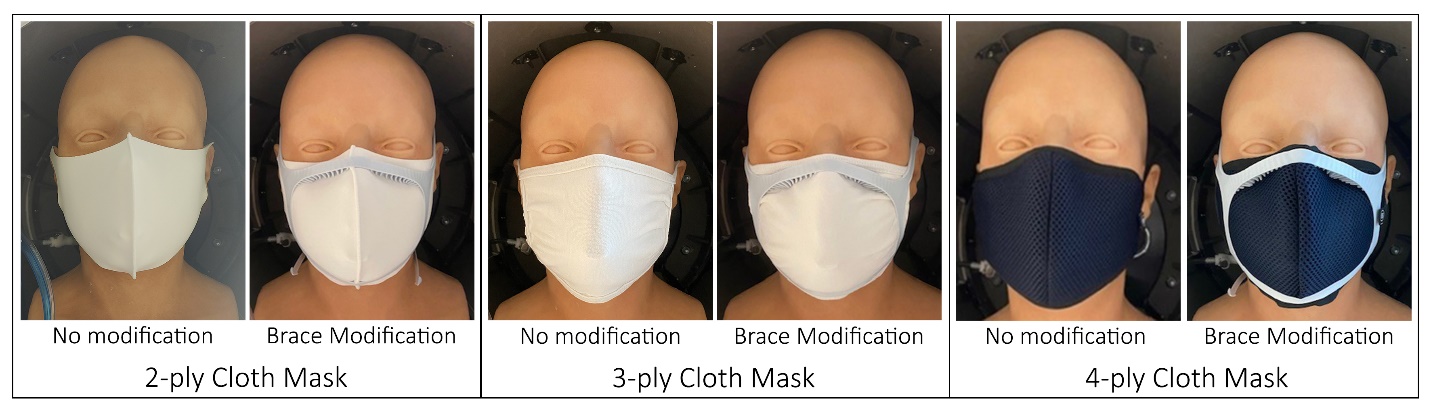

Supplemental Figure S2. Cloth mask fit modifications evaluated during simulated cough and exhalation studies.

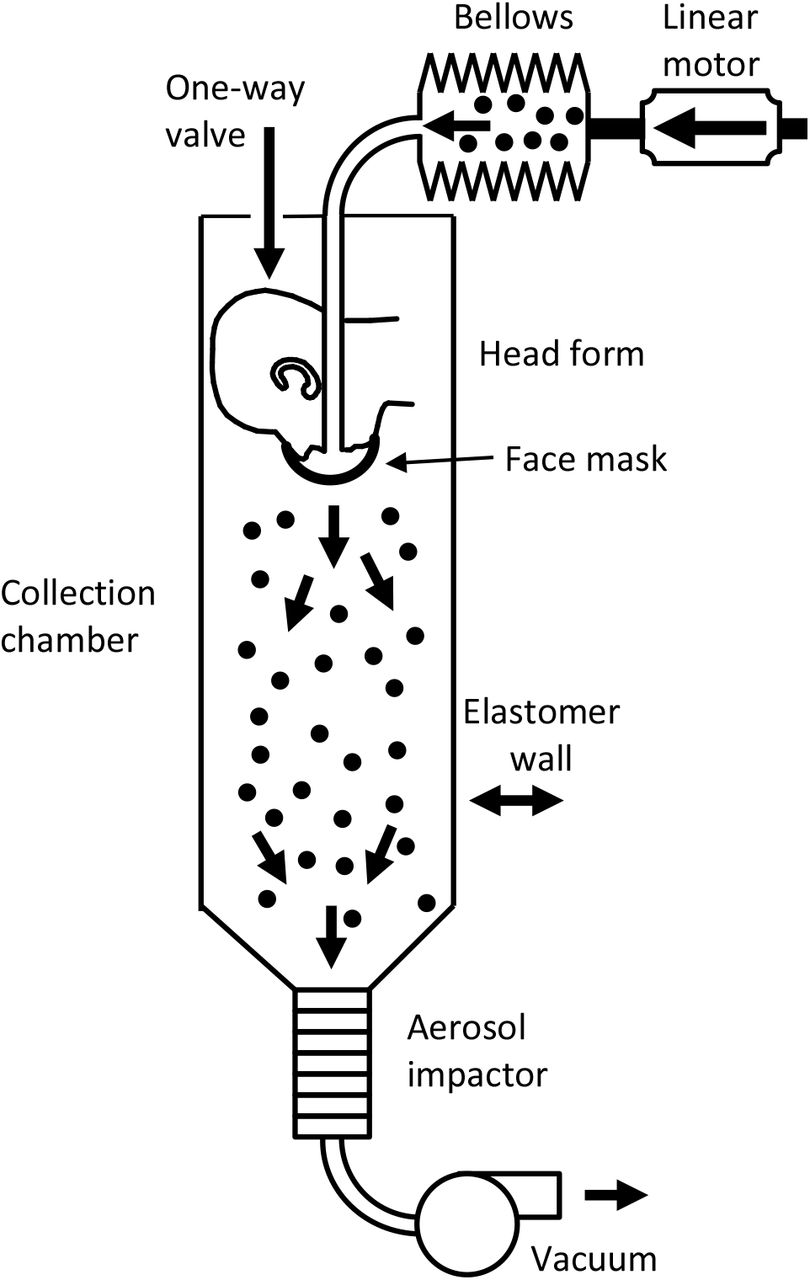

Supplemental Figure S3. Schematic of source control measurement system looking at mask blocking efficacy. The system consists of an aerosol generation system, a bellows and linear motor to produce the simulated coughing and breathing, a pliable skin headform on which the mask is placed, a 136-liter collection chamber into which the aerosol is coughed or exhaled, and a cascade impactor to separate the aerosol particles by size and collect them. The system is described in more detail in Lindsley et al. 2021.^8, 10^

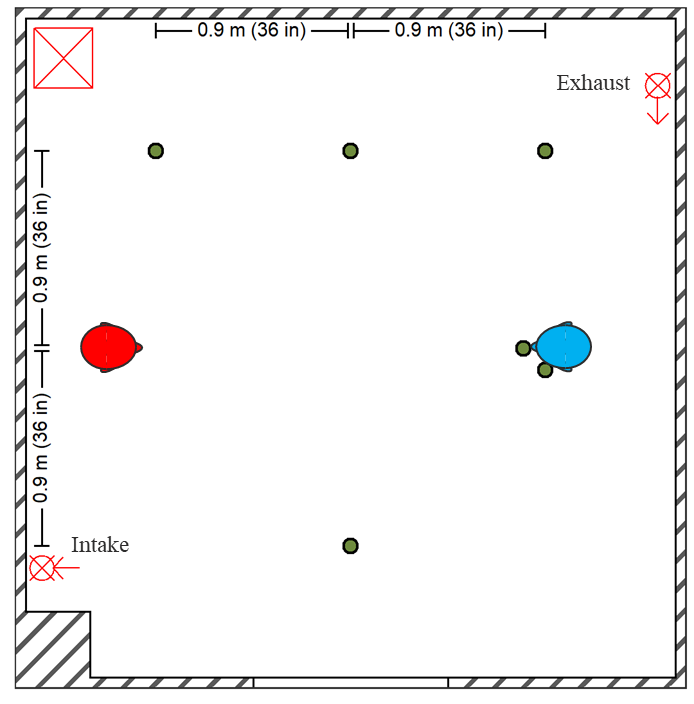

Supplemental Figure S4. Schematic of environment chamber and respiratory simulators used in aerosol exposure masking studies. Diagram of environmental chamber setup showing positions of the aerosol source simulator (red), recipient simulator (blue), and optical particle counters (green dots) for area measurements and personal breathing zone measurements at the mouth and beside the head of the recipient. The HEPA system intake and exhaust are shown with the HEPA filter and blower unit defined by the red square containing an “X”. The system is described by in more detail by Lindsley et al. 2021.^20^
